# Risk of Severe COVID–19 Outcomes Among Patients with Rheumatoid Arthritis in the United States

**DOI:** 10.1101/2021.07.09.21260106

**Authors:** Ching-Yi Chuo, Vince Yau, Sriraman Madhavan, Larry Tsai, Jenny Chia

## Abstract

**Introduction:** Coronavirus disease 2019 (COVID–19) has infected over 22 million individuals worldwide. It remains unclear whether patients with COVID–19 and Rheumatoid Arthritis (RA) experience worse clinical outcomes compared to similar patients with COVID–19 without RA.

**Aim:** The aim of this study is to provide insights on how COVID–19 impacted patients with RA given the nature of the disease and medication used.

**Methods:** RA cases were identified via International Classification of Diseases (ICD) codes and COVID–19 cases by laboratory results in the U.S. based TriNetX network. Patients with COVID–19 and RA were propensity–score matched based on demographics with patients with COVID–19 without RA at a 1:3 ratio. A hospitalized sub-population was defined by procedure codes.

**Results:** We identified 1,014 COVID-19 patients with RA and 3,042 non-RA matches selected from 137,757 patients. The odds of hospitalization (non-RA:23%, RA:24.6%, OR:1.08, 95% CI: 0.88 to 1.33) or mortality (non-RA:5.4%, RA:6%, OR:0.93, 95% CI: 0.65 to 1.34) were not significantly different.

The hospitalized sub-population included 249 patients with COVID-19 and RA and 745 non-RA matches selected from 21,435 patients. The risk of intensive care unit (ICU) admission (non-RA:18.8%, RA:18.1%, OR:0.94, 95% CI: 0.60 to 1.45), and inpatient mortality (non-RA:14.4%, RA:14.5%, OR:0.86, 95% CI: 0.53 to 1.40) were not significantly different.

**Conclusion:** We didn’t find evidence suggesting patients with COVID–19 and RA are more likely to have severe outcomes than patients with COVID–19 without RA.

**Key Messages:**

– Patients with Rheumatoid Arthritis (RA) tend to be older, and often have co-morbidities which could put them at greater risk of severe COVID-19 outcomes.
– This study is one of the largest studies of COVID-19 infected RA populations to date. We did not find increased risk of hospitalization, ICU admission, or mortality among RA patients vs. matched non-RA patients.
– Patients previously exposed to anti-coagulants experienced higher risks of hospitalization and overall mortality. Extra attention is needed for treating such patients.

## Introduction

Coronavirus disease 2019 (COVID-19) has, as of 20 August 2020, infected over 222 million individuals worldwide and caused over 783,000 deaths.^**1**^ Age is the most important risk factor for mortality.^**2**^ Older individuals account for 80% of hospital admissions for COVID-19 infection and mortality risk among hospitalized individuals over 65 years old is 23 times greater than individuals under 65.^**3**^

Increased age is also associated with increased prevalence of comorbidities such as diabetes and hypertension which could contribute to worse outcomes in COVID-19 infected individuals.^**4**^

While several comorbidities such as obesity have been associated with worse outcomes,^**5**^ it is still unclear whether or not patients with COVID-19 and Rheumatoid Arthritis (RA) experience worse clinical outcomes compared to COVID-19 patients without RA. Patients with RA have higher risk of infections due to disease-related immune dysregulation and RA treatments which modulate patient immune responses.^**6 7**^ This has raised patient concerns about whether or not to continue RA treatments during the pandemic.^**8**^

Ironically, several RA medications, rather than being seen as risk factors, have been evaluated as COVID-19 treatments. Several immune modulating RA drugs like dexamethasone have seen widespread use around the world for COVID-19, and others are being evaluated such as interleukin6 (IL-6) inhibitors due to hypotheses that they might reduce risk of severe immune mediated COVID-19 outcomes.^**9**^

In this study, we compare the health outcomes COVID-19 positive patients with and without RA. This analysis will provide insight into how COVID-19 has affected this vulnerable population.

## Methods

### Data Source

The study used the TriNetX (Cambridge, MA) DataWorks network with 41 Healthcare Organizations (HCOs) and 1,016,769 patients across the United States. This COVID-19 specific dataset includes electronic medical records (diagnoses, procedures, medications, and laboratory values) primarily from large academic institutions with better representation of the south (41%, west (23%), midwest (15%) and northwest (17%)).

### Study Population

Patients were required to have confirmed COVID-19 as defined by meeting one of three criteria: 1) Patients with at least one occurrence of International Classification of Diseases, 10th Revision, Clinical Modification (ICD10-CM) code U07.1 or 2) at least one occurrence of ICD10-CM code B97.29 without a negative COVID-19 laboratory test within 7 days or 3) positive antibody or RNA based COVID-19 laboratory test. The study period was between 20 February and 20 August 2020. Index date was defined as the first date fulfilling any of the above definitions.

Patients with COVID-19 were then classified as with RA or without RA. RA was defined as having at least two occurrences of ICD9-CM code 714.0 or corresponding ICD10-CM codes (online supplemental table S1) anytime in their medical records. We also collected information on all RA specific and other treatments such as Disease-Modifying Antirheumatic Drugs (DMARD) such as conventional synthetic (CS), targeted synthetic (TS), and Biologics ([B], steroids, anti-coagulants and antibiotics; online supplemental table S2) taken within the year prior to index date. There are two main populations: 1) individuals who met our COVID-19 definitions and 2) a sub-population of hospitalized patients as defined by procedure codes (table S3).

### Outcomes

The outcomes of interest for all COVID-19 patients were hospitalization and post COVID-19 mortality reported by the HCOs. For the hospitalized sub-population, the outcomes of interest were ICU admission and mortality (online supplemental table S3).

### Statistical Analysis

For each patient-population, we conducted 1:3 propensity score matching using the nearest neighbor method with 0.1 caliper for age, gender, race, and smoking status on their RA status. Two-proportion-z-tests were used to compare the crude proportions of outcomes. Multiple logistic regressions controlling for matching variables, Charlson Comorbidity Index (CCI), Body Mass Index (BMI), and prior medication were used to derive outcome estimates. All analyses were conducted using R (3.5.1)

### Sensitivity Analysis

Sensitivity analyses were conducted by restricting the COVID-19 definition to only lab confirmed cases.

### Ethics

This study used de-identified patient level data from a large electronic medical record database that met Section 164.514 (a) of the HIPAA privacy rule and therefore not considered “research involving human subjects”.

## Results

We identified 138,792 COVID-19 cases and among them, 1,035 were RA patients. Patients with RA and COVID-19 were older (without RA, with RA median age: 44, 63), more likely to be female (without RA, with RA: 55%, 79%), and non-Hispanic (see Table 1). RA patients were also more likely to be smokers (without RA, with RA: 6.5%, 24.5%), with slightly higher BMI and comorbidity burden. RA patients had higher prior-year medication utilization across all treatment classes. However, since the utilization of tsDMARDs was low (n = 93), we did not separately assess tsDAMRD use.

**Table 1.** Demographics, Prior Medication Use and Selected Outcomes Comparison Between Patients with COVID–19 with Rheumatoid Arthritis and Patients with COVID–19 without Rheumatoid Arthritis.

|  | Without RA<br>pre-matched<br>(n = 137,757) | Without RA<br>matched<br>(n=3,042) | With RA<br>(n = 1,035) | All COVID-19<br>(n = 138,792) |
| --- | --- | --- | --- | --- |
| **Age** |  |  |  |  |
| Mean (SD) | 45.0 (19.7) | 61.5 (16.6) | 61.8 (14.6) | 45.2 (19.7) |
| Median [Q1, Q3] | 44.0 [29.0,<br>60.0] | 64.0 [51.0,<br>73.0] | 63.0 [52.0,<br>72.0] | 44.0 [30.0,<br>60.0] |
| Missing | 1,897 (1.4%) |  | 15 (1.4%) | 1,912 (1.4%) |
| **Sex** |  |  |  |  |
| Female | 75,404<br>(54.7%) | 2446 (80.4%) | 816 (78.8%) | 76,220<br>(54.9%) |
| Male | 61,432<br>(44.6%) | 596 (19.6%) | 213 (20.6%) | 61,645<br>(44.4%) |
| Unknown | 921 (0.7%) |  | 6 (0.6%) | 927 (0.7%) |
| **Race** |  |  |  |  |
| American Indian or Alaska Native | 749 (0.5%) | 17 (0.6%) | 2 (0.2%) | 751 (0.5%) |
| Asian | 3,335 (2.4%) | 44 (1.4%) | 18 (1.7%) | 3,353 (2.4%) |
| Black or African American | 32,089<br>(23.3%) | 871 (28.6%) | 286 (27.6%) | 32,375<br>(23.3%) |
| Native Hawaiian or Other Pacific<br>Islander | 306 (0.2%) | 5 (0.2%) | 0 (0%) | 306 (0.2%) |
| White | 68,925<br>(50.0%) | 1508 (49.6%) | 528 (51.0%) | 69,453<br>(50.0%) |
| Unknown | 32,353<br>(23.5%) | 597 (19.6%) | 201 (19.4%) | 32,554<br>(23.5%) |
| **Ethnicity** |  |  |  |  |
| Hispanic or Latino | 25,079<br>(18.2%) | 398 (13.1%) | 124 (12.0%) | 25,203<br>(18.2%) |
| Not Hispanic or Latino | 61,425<br>(44.6%) | 1537 (50.5%) | 541 (52.3%) | 61,966<br>(44.6%) |
| Unknown | 51,253<br>(37.2%) | 1107 (36.4%) | 370 (35.7%) | 51,623<br>(37.2%) |
| **Smoking** |  |  |  |  |
| No | 128,754<br>(93.5%) | 2320 (76.3%) | 781 (75.5%) | 129,535<br>(93.3%) |
| Yes | 9,003 (6.5%) | 722 (23.7%) | 254 (24.5%) | 9257 (6.7%) |
| **BMI (kg/m2)** |  |  |  |  |
| Mean (SD) | 27.8 (5.94) | 28.5 (5.93) | 28.8 (6.02) | 27.8 (5.94) |
| Median [Q1, Q3] | 27.0<br>[23.0, 32.0] | 28.0<br>[24.0, 33.0] | 29.0<br>[25.0, 33.0] | 27.0<br>[23.0, 32.0] |
| Missing | 97,928<br>(71.1%) | 2002 (65.8%) | 490 (47.3%) | 98,418<br>(70.9%) |
| **CCI (weighted score based on** |  |  |  |  |

| **comorbid conditions)** |  |  |  |  |
| --- | --- | --- | --- | --- |
| Mean (SD) | 0.660 (1.68) | 1.39 (2.39) | 3.07 (2.86) | 0.678 (1.71) |
| Median [Q1, Q3] | 0.00 [0.00, 0.00] | 0 [0, 2.00] | 2.00 [1.00, 4.50] | 0.00 [0.00, 0.00] |
| bDMARD= biologics disease–modifying antirheumatic drugs; CCI= Charlson Comorbidity Index; COVID–19= Coronavirus disease 2019; csDMARD= conventional synthetic disease–modifying antirheumatic drugs; ICU= intensive care unit; RA= Rheumatoid Arthritis; SD= standard deviation; tsDMARD= targeted synthetic disease–modifying antirheumatic drugs; Q1= quartile 1; Q3= quartile 3.<br><sup>a</sup> Only among hospitalized patients. |  |  |  |  |

**Table 1 Demographics, Prior Medication Use and Selected Outcomes Comparison Between Patients with COVID–19 with Rheumatoid Arthritis and Patients with COVID–19 without Rheumatoid Arthritis (cont.)**
|  | Without RA<br>(n = 137,757) | Without RA<br>matched<br>(n=3,042) | With RA<br>(n = 1,035) | All COVID-19<br>(n = 138,792) |
| --- | --- | --- | --- | --- |
| **Steroids** |  |  |  |  |
| No | 116,819<br>(84.8%) | 2349 (77.2%) | 497 (48.0%) | 117,316<br>(84.5%) |
| Yes | 20,938<br>(15.2%) | 693 (22.8%) | 538 (52.0%) | 21,476<br>(15.5%) |
| **Anti-coagulants** |  |  |  |  |
| No | 134,593<br>(97.7%) | 2888 (94.9%) | 945 (91.3%) | 135,538<br>(97.7%) |
| Yes | 3,164 (2.3%) | 154 (5.1%) | 90 (8.7%) | 3,254 (2.3%) |
| **Any DMARD** |  |  |  |  |
| No | 136286<br>(98.9%) | 2985 (98.1%) | 601 (58.1%) | 136887<br>(98.6%) |
| Yes | 1471 (1.1%) | 57 (1.9%) | 434 (41.9%) | 1905 (1.4%) |
| **csDMARD** |  |  |  |  |
| No | 136,722<br>(99.2%) | 2997 (98.5%) | 699 (67.5%) | 137,421<br>(99.0%) |
| Yes | 1,035 (0.8%) | 45 (1.5%) | 336 (32.5%) | 1,371 (1.0%) |
| **tsDMARD** |  |  |  |  |
| No | 137,705<br>(100.0%) | 3042 (100%) | 994 (96.0%) | 138,699<br>(99.9%) |
| Yes | 52 (0.0%) | 0 (0%) | 41 (4.0%) | 93 (<0.1%) |
| **bDMARD** |  |  |  |  |
| No | 137,265<br>(99.6%) | 3024 (99.4%) | 844 (81.5%) | 138,109<br>(99.5%) |
| Yes | 492 (0.4%) | 18 (0.6%) | 191 (18.5%) | 683 (0.5%) |
| **Hospitalization** |  |  |  |  |
| No | 116,322<br>(84.4%) | 2342 (77.0%) | 779 (75.3%) | 117,101<br>(84.4%) |
| Yes | 21,435<br>(15.6%) | 700 (23.0%) | 256 (24.7%) | 21,691<br>(15.6%) |
| **ICU<sup>a</sup>** |  |  |  |  |
| No | 17,861<br>(83.3%) | 605 (81.2%) | 210 (82.0%) | 18,071<br>(83.3%) |
| Yes | 3,574<br>(16.7%) | 140 (18.8%) | 46 (18.0%) | 3,620<br>(16.7%) |
| **Inpatient mortality<sup>a</sup>** |  |  |  |  |
| No | 18,949<br>(88.4%) | 638 (85.6%) | 218 (85.2%) | 19,167<br>(88.4%) |
| Yes | 2,486<br>(11.6%) | 107 (14.4%) | 38 (14.8%) | 2,524<br>(11.6%) |
| **Overall mortality** |  |  |  |  |
| No | 134,249<br>(97.5%) | 2,879<br>(94.6%) | 970 (93.7%) | 135,219<br>(97.4%) |
| Yes | 3,508 (2.5%) | 163 (5.4%) | 65 (6.3%) | 3,573 (2.6%) |
| bDMARD = biologics disease-modifying antirheumatic drugs; CCI = Charlson Comorbidity Index;<br>COVID-19 = Coronavirus disease 2019; csDMARD = conventional synthetic disease-modifying<br>antirheumatic drugs; ICU = intensive care unit; RA = Rheumatoid Arthritis; SD = standard deviation;<br>tsDMARD = targeted synthetic disease-modifying antirheumatic drugs; Q1 = quartile 1; Q3 = quartile 3.<br><sup>a</sup> Only among hospitalized patients. |  |  |  |  |

Comparing the pre-matched crude proportions of the outcomes between COVID-19 patients with RA and (n=1,035) and without RA (n=137,757), the former group had a significantly higher hospitalization rate (24.7% vs. 15.6%, p <0.05) and mortality (6.3% vs. 2.5%, p <0.05). Once hospitalized, the patients with and without RA had similar likelihoods of transferring to the ICU (with RA 18.0%, without RA 16.7%, p = 0.64) and inpatient mortality (with RA 14.8%, without RA 11.6%, p = 0.13).

We matched 3,042 non-RA COVID-19 patients to 1,014 RA COVID-19 patients and 745 hospitalized non-RA COVID-19 patients to 249 hospitalized RA COVID-19 patients. All matching variables were well balanced post-match (Figure S1/S2). When comparing non-hospitalized patients with RA and matched patients without RA, we found no significant difference in the odds of hospitalization and mortality (hospitalization odds ratio [OR] 1.08, 95% confidence interval [CI]: [0.88,1.33]), mortality (OR 0.93, 95% [CI]: [0.65,1.34]). For matched hospitalized COVID-19 patients with RA (n=249) and patients without RA (n=745), we did not find significant differences in odds of transferring to ICU, or death during hospitalization (ICU: OR 0.94, 95% [CI]: [0.60,1.45], inpatient mortality: OR 0.86, 95% [CI]: [0.53,1.40]) (see Table 2). Treatments up to a year prior to the index date were also evaluated (Table 3). We found no significant effects except for anti-coagulants on the risk of hospitalization (OR 1.49, 95% [CI]: [1.10,2.02]) and overall mortality (OR 1.62, 95% [CI]: [1.04, 2.51]).

**Table 2.** Outcomes Among Patients with Rheumatoid Arthritis with COVID–19 and Matched Controls.

| a. Outcomes all COVID-19 patients |  |  |  |  |  |
| --- | --- | --- | --- | --- | --- |
|  | Matched Without RA<br>(n=3042) | With RA<br>(n=1014) | Overall<br>(n=4056) | Crude OR<br>(95% CI) | Adjusted OR<sup>a</sup><br>(95% CI) |
| **Hospitalization** |  |  |  |  |  |
| No | 2,342<br>(77.0%) | 765<br>(75.4%) | 3,107<br>(76.6%) | 1.78<br>(1.55, 2.06) | 1.08<br>(0.88, 1.33) |
| Yes | 700<br>(23.0%) | 249<br>(24.6%) | 949<br>(23.4%) |  |  |
| **Mortality** |  |  |  |  |  |
| No | 2,879<br>(94.6%) | 953<br>(94.0%) | 3,832<br>(94.5%) | 2.56<br>(1.99, 3.30) | 0.93<br>(0.65, 1.34) |
| Yes | 163 (5.4%) | 61<br>(6.0%) | 224<br>(5.5%) |  |  |
| b. Outcomes among hospitalized COVID-19 patients |  |  |  |  |  |
|  | Matched Without RA<br>(n=745) | With RA<br>(n=249) | Overall<br>(n=994) | Crude OR<br>(95% CI) | Adjusted OR<sup>a</sup><br>(95% CI) |
| **ICU** |  |  |  |  |  |
| No | 605<br>(81.2%) | 204<br>(81.9%) | 809<br>(81.4%) | 1.09<br>(0.79 1.51) | 0.94<br>(0.60, 1.45) |
| Yes | 140<br>(18.8%) | 45<br>(18.1%) | 185<br>(18.6%) |  |  |
| **Inpatient** |  |  |  |  |  |

| mortality |  |  |  |  |  |
| --- | --- | --- | --- | --- | --- |
| No | 638<br>(85.6%) | 213<br>(85.5%) | 851<br>(85.6%) | 1.33<br>(0.94, 1.88) | 0.86<br>(0.53, 1.40) |
| Yes | 107<br>(14.4%) | 36<br>(14.5%) | 143<br>(14.4%) |  |  |

CI = confidence interval; OR=odds ratio; RA=Rheumatoid Arthritis.
<sup>a</sup> Results are based on multiple logistic regression adjusting for age, race/ethnicity, sex, body mass index (BMI), comorbidities, and pre-index medication such as steroids, anti-coagulants, antibiotics, csDMARD and bDMARD.

**Table 3.** The effects of treatment on different outcomes evaluated.

|  | Outcomes among all COVID-19 patients<br>OR (95% CI) |  | Outcomes among all Hospitalized COVID-19 patients<br>OR (95% CI) |  |
| --- | --- | --- | --- | --- |
| Treatment | Hospitalization | OS | ICU | Inpatient mortality |
| Steroids | 0.83 (0.68, 1.01) | 0.99 (0.70, 1.40) | 0.93 (0.60, 1.44) | 0.85 (0.53, 1.37) |
| Anti-coagulants | 1.49 (1.10, 2.02) | 1.62 (1.04, 2.51) | 1.68 (0.97, 2.94) | 1.44 (0.78, 2.68) |
| csDMARD | 0.76 (0.55, 1.04) | 0.66 (0.36, 1.22) | 0.73 (0.35, 1.52) | 0.76 (0.33, 1.76) |
| bDMARD | 0.69 (0.45, 1.05) | 1.19 (0.58, 2.42) | 1.87 (0.78, 4.49) | 2.42 (0.93, 6.29) |

CI = confidence interval; OR = odds ratio; RA = Rheumatoid Arthritis; OS = overall survival. Results are based on multiple logistic regression adjusting for sex, age, race/ethnicity, body mass index (BMI), and comorbidities.

Our sensitivity analyses looking at laboratory-confirmed COVID-19 cases reduced our sample size by 32%. Results remained unchanged except that confidence intervals were wider (table S4-S6).

## Discussion

It has been hypothesized that persons using immunosuppressive medications might be at higher risk of severe COVID-19 outcomes due to a compromised immune system and inability to suppress early viral replication. However, we did not find evidence to suggest patients with COVID-19 and RA are more likely to have severe outcomes after matching. We also did not find any effect of pre-COVID-19 DMARD treatments on the risk of COVID-19 related outcomes.

There is a growing body of conflicting literature examining COVID-19 outcomes among RA patients. While some studies reported increased risk of hospitalization^15^ and death^16^ among combined populations of patients with inflammatory conditions, specific analyses focusing on RA populations have suggested that there is no increased risk of hospitalization.^7 17^ However, when examining RA treatments, some studies suggest a possible protective effect of b/tsDMARDs^7^ and anti-TNF therapy^7 15^ on risk of hospitalization, while increased risk of hospitalization was reported for JAK inhibitor users^15^. Directionally, findings from our paper also suggested similar results, with a non-statistically significant reduced risk of hospitalization among bDMARD users. Subsequent analysis of the hospitalized patients showed non-statistically significant elevated risks of ICU stay and mortality. Notably, in our sensitivity analyses using only lab confirmed COVID-19 cases, the directional association showing reduced risk of hospitalization remained for all COVID-19 patients, while among hospitalized patients, the risk of ICU and mortality was no longer elevated.

When characterizing patients with and without severe outcomes, we found older age, male gender, smokers, and higher CCI are associated with severe outcomes (online supplemental table S7-S8). Patients utilizing anti-coagulants in the pre-COVID-19 period had significantly increased risk of hospitalization and mortality (Table 3). Similar, non-significant results were found among hospitalized patients (online supplemental table S9-S10). The effect of anti-coagulant use was the same across populations with or without RA.

COVID-19 is known to be associated with abnormal coagulation, specifically higher clot formation^**12**^ and subsequent increased risk of mortality.^**13 14**^ Patients using anti-coagulants may have comorbidities that predispose them to worse outcomes from COVID-19 (cardiovascular disease, malignancy, or another hypercoagulable state) despite the potential protective effect of the anti-coagulant itself. Alternatively, the anti-coagulants themselves could paradoxically be causing worse COVID-19 outcomes, perhaps by increasing the risk of bleeding complications. Distinguishing between these hypotheses could have important implications for the treatment of patients with COVID-19; however, further study is required.

Although the scope of this study is limited to information captured by participating HCOs and might not be reflective of patients’ complete medical encounters, medication use and detailed disease conditions such as disease activities, this is the largest published study examining the risk of severe outcomes among patients with COVID-19 and RA to date. Worse COVID-19 outcomes were not seen among patients with RA, regardless of historical RA therapy. These findings suggest that asymptomatic patients with RA should stay on their routine RA therapy, though it is still unclear whether or not RA or other medications should be discontinued after COVID-19 infection.

## Supporting information

All supplemental tables

## Data Availability

We are not providing the patient-level data per TriNetX's policy. TriNetX maintains processes and procedures to respond to questions about the data. If editors or reviewers have questions about TriNetX data, TriNetX provides details on the data provenance and/or the necessary access to audit the processes. If the validity of the data is questioned further, TriNetX would make the data available for a third party audit.

## Acknowledgements

**N/A**

