## Supplementary material for "Risk of Severe COVID–19 Outcomes Among Patients with Rheumatoid Arthritis in the United States": All supplemental tables

**Table S1	Rheumatoid Arthritis Diagnostic Codes**

| ICD−10 Code | ICD−10 Description | ICD−9 Code | ICD−10 Description |
| --- | --- | --- | --- |
| M0570 | RA with rheumatoid factor of unspecified site without organ or systems involvement | 7140 | RA |
| M05711 | RA with rheumatoid factor of right shoulder without organ or systems involvement | 7140 | RA |
| M05712 | RA with rheumatoid factor of left shoulder without organ or systems involvement | 7140 | RA |
| M05719 | RA with rheumatoid factor of unspecified shoulder without organ or systems involvement | 7140 | RA |
| M05721 | RA with rheumatoid factor of right elbow without organ or systems involvement | 7140 | RA |
| M05722 | RA with rheumatoid factor of left elbow without organ or systems involvement | 7140 | RA |
| M05729 | RA with rheumatoid factor of unspecified elbow without organ or systems involvement | 7140 | RA |
| M05731 | RA with rheumatoid factor of right wrist without organ or systems involvement | 7140 | RA |
| M05732 | RA with rheumatoid factor of left wrist without organ or systems involvement | 7140 | RA |
| M05739 | RA with rheumatoid factor of unspecified wrist without organ or systems involvement | 7140 | RA |
| M05741 | RA with rheumatoid factor of right hand without organ or systems involvement | 7140 | RA |
| M05742 | RA with rheumatoid factor of left hand without organ or systems involvement | 7140 | RA |
| M05749 | RA with rheumatoid factor of unspecified hand without organ or systems involvement | 7140 | RA |
| M05751 | RA with rheumatoid factor of right hip without organ or systems involvement | 7140 | RA |
| M05752 | RA with rheumatoid factor of left hip without organ or systems involvement | 7140 | RA |
| M05759 | RA with rheumatoid factor of unspecified hip without organ or systems involvement | 7140 | RA |
| M05761 | RA with rheumatoid factor of right knee without organ or systems involvement | 7140 | RA |
| M05762 | RA with rheumatoid factor of left knee without organ or systems involvement | 7140 | RA |
| M05769 | RA with rheumatoid factor of unspecified knee without organ or systems involvement | 7140 | RA |
| M05771 | RA with rheumatoid factor of right ankle and foot without organ or systems involvement | 7140 | RA |
| M05772 | RA with rheumatoid factor of left ankle and foot without organ or systems involvement | 7140 | RA |
| M05779 | RA with rheumatoid factor of unspecified ankle and foot without organ or systems involvement. | 7140 | RA |
| M0579 | RA with rheumatoid factor of multiple sites without organ or systems involvement | 7140 | RA |
| ICD−9 = International Classification of Diseases, 9^th^ Revision; ICD−10 = International Classification of Diseases, 10^th^ Revision; RA = Rheumatoid Arthritis | | | |

**Table S1	Rheumatoid Arthritis Diagnostic Codes (cont.)**

| ICD−10 Code | ICD−10 Description | ICD−9 Code | ICD−10 description |
| --- | --- | --- | --- |
| M0580 | Other RA with rheumatoid factor of unspecified site | 7140 | RA |
| M05811 | Other RA with rheumatoid factor of right shoulder | 7140 | RA |
| M05812 | Other RA with rheumatoid factor of left shoulder | 7140 | RA |
| M05819 | Other RA with rheumatoid factor of unspecified shoulder | 7140 | RA |
| M05821 | Other RA with rheumatoid factor of right elbow | 7140 | RA |
| M05822 | Other RA with rheumatoid factor of left elbow | 7140 | RA |
| M05829 | Other RA with rheumatoid factor of unspecified elbow | 7140 | RA |
| M05831 | Other RA with rheumatoid factor of right wrist | 7140 | RA |
| M05832 | Other RA with rheumatoid factor of left wrist | 7140 | RA |
| M05839 | Other RA with rheumatoid factor of unspecified wrist | 7140 | RA |
| M05841 | Other RA with rheumatoid factor of right hand | 7140 | RA |
| M05842 | Other RA with rheumatoid factor of left hand | 7140 | RA |
| M05849 | Other RA with rheumatoid factor of unspecified hand | 7140 | RA |
| M05851 | Other RA with rheumatoid factor of right hip | 7140 | RA |
| M05852 | Other RA with rheumatoid factor of left hip | 7140 | RA |
| M05859 | Other RA with rheumatoid factor of unspecified hip | 7140 | RA |
| M05861 | Other RA with rheumatoid factor of right knee | 7140 | RA |
| M05862 | Other RA with rheumatoid factor of left knee | 7140 | RA |
| M05869 | Other RA with rheumatoid factor of unspecified knee | 7140 | RA |
| M05871 | Other RA with rheumatoid factor of right ankle and foot | 7140 | RA |
| M05872 | Other RA with rheumatoid factor of left ankle and foot | 7140 | RA |
| M05879 | Other RA with rheumatoid factor of unspecified ankle and foot | 7140 | RA |
| M0589 | Other RA with rheumatoid factor of multiple sites | 7140 | RA |
| M059 | RA with rheumatoid factor, unspecified | 7140 | RA |
| M0600 | RA without rheumatoid factor, unspecified site | 7140 | RA |
| M06011 | RA without rheumatoid factor, right shoulder | 7140 | RA |
| M06012 | RA without rheumatoid factor, left shoulder | 7140 | RA |
| M06019 | RA without rheumatoid factor, unspecified shoulder | 7140 | RA |
| M06021 | RA without rheumatoid factor, right elbow | 7140 | RA |
| M06022 | RA without rheumatoid factor, left elbow | 7140 | RA |
| M06029 | RA without rheumatoid factor, unspecified elbow | 7140 | RA |
| M06031 | RA without rheumatoid factor, right wrist | 7140 | RA |
| M06032 | RA without rheumatoid factor, left wrist | 7140 | RA |
| M06039 | RA without rheumatoid factor, unspecified wrist | 7140 | RA |
| M06041 | RA without rheumatoid factor, right hand | 7140 | RA |
| M06042 | RA without rheumatoid factor, left hand | 7140 | RA |
| M06049 | RA without rheumatoid factor, unspecified hand | 7140 | RA |
| M06051 | RA without rheumatoid factor, right hip | 7140 | RA |
| M06052 | RA without rheumatoid factor, left hip | 7140 | RA |
| ICD−9 = International Classification of Diseases, 9^th^ Revision; ICD−10 = International Classification of Diseases, 10^th^ Revision; RA = Rheumatoid Arthritis | | | |

**Table S1	Rheumatoid Arthritis Diagnostic Codes (cont.)**

| ICD−10 Code | ICD−10 Description | ICD−9 Code | ICD−10 description |
| --- | --- | --- | --- |
| M06059 | RA without rheumatoid factor, unspecified hip | 7140 | RA |
| M06061 | RA without rheumatoid factor, right knee | 7140 | RA |
| M06062 | RA without rheumatoid factor, left knee | 7140 | RA |
| M06069 | RA without rheumatoid factor, unspecified knee | 7140 | RA |
| M06071 | RA without rheumatoid factor, right ankle and foot | 7140 | RA |
| M06072 | RA without rheumatoid factor, left ankle and foot | 7140 | RA |
| M06079 | RA without rheumatoid factor, unspecified ankle and foot | 7140 | RA |
| M0608 | RA without rheumatoid factor, vertebrae | 7140 | RA |
| M0609 | RA without rheumatoid factor, multiple sites | 7140 | RA |
| M0620 | Rheumatoid bursitis, unspecified site | 7140 | RA |
| M06211 | Rheumatoid bursitis, right shoulder | 7140 | RA |
| M06212 | Rheumatoid bursitis, left shoulder | 7140 | RA |
| M06219 | Rheumatoid bursitis, unspecified shoulder | 7140 | RA |
| M06221 | Rheumatoid bursitis, right elbow | 7140 | RA |
| M06222 | Rheumatoid bursitis, left elbow | 7140 | RA |
| M06229 | Rheumatoid bursitis, unspecified elbow | 7140 | RA |
| M06231 | Rheumatoid bursitis, right wrist | 7140 | RA |
| M06232 | Rheumatoid bursitis, left wrist | 7140 | RA |
| M06239 | Rheumatoid bursitis, unspecified wrist | 7140 | RA |
| M06241 | Rheumatoid bursitis, right hand | 7140 | RA |
| M06242 | Rheumatoid bursitis, left hand | 7140 | RA |
| M06249 | Rheumatoid bursitis, unspecified hand | 7140 | RA |
| M06251 | Rheumatoid bursitis, right hip | 7140 | RA |
| M06252 | Rheumatoid bursitis, left hip | 7140 | RA |
| M06259 | Rheumatoid bursitis, unspecified hip | 7140 | RA |
| M06261 | Rheumatoid bursitis, right knee | 7140 | RA |
| M06262 | Rheumatoid bursitis, left knee | 7140 | RA |
| M06269 | Rheumatoid bursitis, unspecified knee | 7140 | RA |
| M06271 | Rheumatoid bursitis, right ankle and foot | 7140 | RA |
| M06272 | Rheumatoid bursitis, left ankle and foot | 7140 | RA |
| M06279 | Rheumatoid bursitis, unspecified ankle and foot | 7140 | RA |
| M0628 | Rheumatoid bursitis, vertebrae | 7140 | RA |
| M0629 | Rheumatoid bursitis, multiple sites | 7140 | RA |
| M0630 | Rheumatoid nodule, unspecified site | 7140 | RA |
| M06311 | Rheumatoid nodule, right shoulder | 7140 | RA |
| M06312 | Rheumatoid nodule, left shoulder | 7140 | RA |
| M06319 | Rheumatoid nodule, unspecified shoulder | 7140 | RA |
| M06321 | Rheumatoid nodule, right elbow | 7140 | RA |
| M06322 | Rheumatoid nodule, left elbow | 7140 | RA |
| M06329 | Rheumatoid nodule, unspecified elbow | 7140 | RA |
| M06331 | Rheumatoid nodule, right wrist | 7140 | RA |
| M06332 | Rheumatoid nodule, left wrist | 7140 | RA |
| ICD−9 = International Classification of Diseases, 9^th^ Revision; ICD−10 = International Classification of Diseases, 10^th^ Revision; RA = Rheumatoid Arthritis | | | |

**Table S1	Rheumatoid Arthritis Diagnostic Codes (cont.)**

| ICD−10 Code | ICD−10 Description | ICD−9 Code | ICD−10 description |
| --- | --- | --- | --- |
| M06339 | Rheumatoid nodule, unspecified wrist | 7140 | RA |
| M06341 | Rheumatoid nodule, right hand | 7140 | RA |
| M06342 | Rheumatoid nodule, left hand | 7140 | RA |
| M06349 | Rheumatoid nodule, unspecified hand | 7140 | RA |
| M06351 | Rheumatoid nodule, right hip | 7140 | RA |
| M06352 | Rheumatoid nodule, left hip | 7140 | RA |
| M06359 | Rheumatoid nodule, unspecified hip | 7140 | RA |
| M06361 | Rheumatoid nodule, right knee | 7140 | RA |
| M06362 | Rheumatoid nodule, left knee | 7140 | RA |
| M06369 | Rheumatoid nodule, unspecified knee | 7140 | RA |
| M06371 | Rheumatoid nodule, right ankle and foot | 7140 | RA |
| M06372 | Rheumatoid nodule, left ankle and foot | 7140 | RA |
| M06379 | Rheumatoid nodule, unspecified ankle and foot | 7140 | RA |
| M0638 | Rheumatoid nodule, vertebrae | 7140 | RA |
| M0639 | Rheumatoid nodule, multiple sites | 7140 | RA |
| M0680 | Other specified RA, unspecified site | 7140 | RA |
| M06811 | Other specified RA, right shoulder | 7140 | RA |
| M06812 | Other specified RA, left shoulder | 7140 | RA |
| M06819 | Other specified RA, unspecified shoulder | 7140 | RA |
| M06821 | Other specified RA, right elbow | 7140 | RA |
| M06822 | Other specified RA, left elbow | 7140 | RA |
| M06829 | Other specified RA, unspecified elbow | 7140 | RA |
| M06831 | Other specified RA, right wrist | 7140 | RA |
| M06832 | Other specified RA, left wrist | 7140 | RA |
| M06839 | Other specified RA, unspecified wrist | 7140 | RA |
| M06841 | Other specified RA, right hand | 7140 | RA |
| M06842 | Other specified RA, left hand | 7140 | RA |
| M06849 | Other specified RA, unspecified hand | 7140 | RA |
| M06851 | Other specified RA, right hip | 7140 | RA |
| M06852 | Other specified RA, left hip | 7140 | RA |
| M06859 | Other specified RA, unspecified hip | 7140 | RA |
| M06861 | Other specified RA, right knee | 7140 | RA |
| M06862 | Other specified RA, left knee | 7140 | RA |
| M06869 | Other specified RA, unspecified knee | 7140 | RA |
| M06871 | Other specified RA, right ankle and foot | 7140 | RA |
| M06872 | Other specified RA, left ankle and foot | 7140 | RA |
| M06879 | Other specified RA, unspecified ankle and foot | 7140 | RA |
| M0688 | Other specified RA, vertebrae | 7140 | RA |
| M0689 | Other specified RA, multiple sites | 7140 | RA |
| M069 | RA, unspecified | 7140 | RA |
| M0540 | Rheumatoid myopathy with RA of unspecified site | 7140 | RA |
| M05411 | Rheumatoid myopathy with RA of right shoulder | 7140 | RA |
| M05412 | Rheumatoid myopathy with RA of left shoulder | 7140 | RA |
| ICD−9 = International Classification of Diseases, 9^th^ Revision; ICD−10 = International Classification of Diseases, 10^th^ Revision; RA = Rheumatoid Arthritis | | | |

**Table S1	Rheumatoid Arthritis Diagnostic Codes (cont.)**

| ICD−10 Code | ICD−10 Description | ICD−9 Code | ICD−10 description |
| --- | --- | --- | --- |
| M05419 | Rheumatoid myopathy with RA of unspecified shoulder | 7140 | RA |
| M05421 | Rheumatoid myopathy with RA of right elbow | 7140 | RA |
| M05422 | Rheumatoid myopathy with RA of left elbow | 7140 | RA |
| M05429 | Rheumatoid myopathy with RA of unspecified elbow | 7140 | RA |
| M05431 | Rheumatoid myopathy with RA of right wrist | 7140 | RA |
| M05432 | Rheumatoid myopathy with RA of left wrist | 7140 | RA |
| M05439 | Rheumatoid myopathy with RA of unspecified wrist | 7140 | RA |
| M05441 | Rheumatoid myopathy with RA of right hand | 7140 | RA |
| M05442 | Rheumatoid myopathy with RA of left hand | 7140 | RA |
| M05449 | Rheumatoid myopathy with RA of unspecified hand | 7140 | RA |
| M05451 | Rheumatoid myopathy with RA of right hip | 7140 | RA |
| M05452 | Rheumatoid myopathy with RA of left hip | 7140 | RA |
| M05459 | Rheumatoid myopathy with RA of unspecified hip | 7140 | RA |
| M05461 | Rheumatoid myopathy with RA of right knee | 7140 | RA |
| M05462 | Rheumatoid myopathy with RA of left knee | 7140 | RA |
| M05469 | Rheumatoid myopathy with RA of unspecified knee | 7140 | RA |
| M05471 | Rheumatoid myopathy with RA of right ankle and foot | 7140 | RA |
| M05472 | Rheumatoid myopathy with RA of left ankle and foot | 7140 | RA |
| M05479 | Rheumatoid myopathy with RA of unspecified ankle and foot | 7140 | RA |
| M0549 | Rheumatoid myopathy with RA of multiple sites | 7140 | RA |
| M0550 | Rheumatoid polyneuropathy with RA of unspecified site | 7140 | RA |
| M05511 | Rheumatoid polyneuropathy with RA of right shoulder | 7140 | RA |
| M05512 | Rheumatoid polyneuropathy with RA of left shoulder | 7140 | RA |
| M05519 | Rheumatoid polyneuropathy with RA of unspecified shoulder | 7140 | RA |
| ICD−9 = International Classification of Diseases, 9^th^ Revision; ICD−10 = International Classification of Diseases, 10^th^ Revision; RA = Rheumatoid Arthritis | | | |

**Table S1	Rheumatoid Arthritis Diagnostic Codes (cont.)**

| ICD−10 Code | ICD−10 Description | ICD−9 Code | ICD−10 description |
| --- | --- | --- | --- |
| M05521 | Rheumatoid polyneuropathy with RA of right elbow | 7140 | RA |
| M05522 | Rheumatoid polyneuropathy with RA of left elbow | 7140 | RA |
| M05529 | Rheumatoid polyneuropathy with RA of unspecified elbow | 7140 | RA |
| M05531 | Rheumatoid polyneuropathy with RA of right wrist | 7140 | RA |
| M05532 | Rheumatoid polyneuropathy with RA of left wrist | 7140 | RA |
| M05539 | Rheumatoid polyneuropathy with RA of unspecified wrist | 7140 | RA |
| M05541 | Rheumatoid polyneuropathy with RA of right hand | 7140 | RA |
| M05542 | Rheumatoid polyneuropathy with RA of left hand | 7140 | RA |
| M05549 | Rheumatoid polyneuropathy with RA of unspecified hand | 7140 | RA |
| M05551 | Rheumatoid polyneuropathy with RA of right hip | 7140 | RA |
| M05552 | Rheumatoid polyneuropathy with RA of left hip | 7140 | RA |
| M05559 | Rheumatoid polyneuropathy with RA of unspecified hip | 7140 | RA |
| M05561 | Rheumatoid polyneuropathy with RA of right knee | 7140 | RA |
| M05562 | Rheumatoid polyneuropathy with RA of left knee | 7140 | RA |
| M05569 | Rheumatoid polyneuropathy with RA of unspecified knee | 7140 | RA |
| M05571 | Rheumatoid polyneuropathy with RA of right ankle and foot | 7140 | RA |
| M05572 | Rheumatoid polyneuropathy with RA of left ankle and foot | 7140 | RA |
| M05579 | Rheumatoid polyneuropathy with RA of unspecified ankle and foot | 7140 | RA |
| M0559 | Rheumatoid polyneuropathy with RA of multiple sites | 7140 | RA |
| ICD−9 = International Classification of Diseases, 9^th^ Revision; ICD−10 = International Classification of Diseases, 10^th^ Revision; RA = Rheumatoid Arthritis | | | |

**Table S2	List of Medications Based on Treatment Group**

| Treatment Group | Medication |
| --- | --- |
| Antibiotics | Oxytetracycline hcl, tetracycline, clarithromycin, minocycline hcl, tigecycline, eravacycline, omadacycline, chloramphenicol, ampicillin sodium, piperacillin sodium, ticarcillin disodium, oxacillin sodium, nafcillin sodium, cefazolin, cefuroxime, cefotetan disodium, cefotaxime sodium, ceftazidime, ceftriaxone sodium, ceftizoxime sodium, cefoperazone sodium, cefepime hcl, aztreonam, meropenem, ertapenem sodium, doripenem, ceftaroline fosamil, sulfamethoxazole, erythromycin, azithromycin, clindamycin, lincomycin hcl, quinupristin, streptomycin, tobramycin, garamycin, kanamycin, amikacin, ciprofloxacin, ofloxacin, ciprofloxacin, alatrofloxacin, ofloxacin, ciprofloxacin, moxifloxacin, gatifloxacin, delafloxacin, vancomycin, telavancin, dalbavancin, metronidazole, spectinomycin dihydrochloride, linezolid, daptomycin, tedizolid phosphate |
| Steroids | cortisone acetate, dexamethasone, hydrocortisone, methylprednisolone, prednisolone, prednisone |
| Anti−coagulants | Warfarin, venus thromboembolism (VTE), argatroban, heparin, alteplase, fondaparinux sodium, dalteparin sodium, tinzaparin sodium, tinzaparin, iloprost, bivalirudin, eptifiban hcl, antithrombin III, Treprostinil, reteplase, abciximab, urokinease, epoprostenol, treprostinol, caplacizumab, Tenecteplase, anistreplase, cangrelor, drotrecogin alpha |
| csDMARD | Chloroquine/hydroxychloroquine, methotrexate (MTX), sulfasalazine (SSZ) |
| tsDMARD | apremilast, baricitinib, filgotinib, tofacitinib |
| bDMARD | Abatacept, adalimumab, anakinra, certolizumab, etanercept, golimumab, infliximab, ixekizumab, natalizumab, rituximab, secukinumab, tocilizumab, ustekinumab, vedolizumab |
| bDMARD = biologics disease−modifying antirheumatic drugs; csDMARD = conventional synthetic disease−modifying antirheumatic drugs; tsDMARD = targeted synthetic disease−modifying antirheumatic drugs. | |

**Table S3	Procedure Codes for Defining Outcomes**

| Outcome | Procedure codes |
| --- | --- |
| Hospitalization | CPT: 99221, 99222, 99223, 99231, 99232, 99233, 99234, 99235, 99236, 99238, 99239, 99291, 99292  Any ICD−10-PCS codes |
| ICU | A subset of hospitalization with CPT: 99291, 99292 |
| Outcome | Procedure codes |
| ICD−10-PCS = International Classification of Diseases, 10^th^ Revision Procedure Coding System; ICU = intensive care unit. | |

**Table S4**	**Sensitivity Analysis Among Laboratory−Confirmed Patients with Rheumatoid Arthritis**

|  | Matched Without Rheumatoid Arthritis | With Rheumatoid Arthritis (n = 504) | Matched Without Rheumatoid Arthritis | Adjusted OR ^a^ (95% CI) |
| --- | --- | --- | --- | --- |
| **Hospitalization** |  |  |  |  |
| No | 1,461 (75.3%) | 369 (73.2%) | 1,830 (74.8%) | 1.13 (0.85, 1.50) |
| Yes | 480 (24.7%) | 135 (26.8%) | 615 (25.2%) |  |
| **Death** |  |  |  |  |
| No | 1,844 (95.0%) | 472 (93.7%) | 2,316 (93.7%) | 1.00 (0.61, 1.66) |
| Yes | 97 (5.0%) | 32 (6.3%) | 129 (5.3%) |  |
| CI = confidence interval; OR = odds ratio; RA = Rheumatoid Arthritis.  ^a^ Results are based on multiple logistic regression adjusting for age, race/ethnicity, sex, body mass index (BMI), comorbidities, and pre−index medication such as steroids, anti-coagulants, antibiotics, csDMARD and bDMARD. | | | | |

**Table S5**	**Sensitivity Analysis Among Hospitalized Laboratory−Confirmed Patients with Rheumatoid Arthritis**

|  | Matched Without Rheumatoid Arthritis (n = 402) | With Rheumatoid Arthritis (n = 135) | Overall (n = 537) | Adjusted OR ^a^ (95% CI) |
| --- | --- | --- | --- | --- |
| **ICU** |  |  |  |  |
| No | 339 (84.3%) | 112 (83.0%) | 451 (84.0%) | 1.10 (0.58, 2.08) |
| Yes | 63 (15.7%) | 23 (17.0%) | 86 (16.0%) |  |
| **Death** |  |  |  |  |
| No | 347 (86.3%) | 119 (88.1%) | 466 (86.8%) | 0.85 (0.43, 1.69) |
| xs | 55 (13.7%) | 16 (11.9%) | 71 (13.2%) |  |
| CI = confidence interval; OR = odds ratio; RA = Rheumatoid Arthritis.  ^a^ Results are based on multiple logistic regression adjusting for age, race/ethnicity, sex, body mass index (BMI), comorbidities, and pre−index medication such as steroids, anti-coagulants, antibiotics, csDMARD and bDMARD. | | | | |

**Table S6	Sensitivity analysis of Treatment Effects Among Hospitalized Laboratory−Confirmed Patients with Rheumatoid Arthritis**

|  | Outcomes among all COVID−19 patients OR (95% CI) | | Outcomes among all Hospitalized COVID−19 patients OR (95% CI) | |
| --- | --- | --- | --- | --- |
| Treatment | Hospitalization | OS | ICU | Inpatient Mortality |
| Steroids | 0.82 (0.64, 1.07) | 1.19 (0.75, 1.88) | 0.77 (0.41, 1.45) | 0.83 (0.42, 1.64) |
| Anti−coagulants | 1.25 (0.84, 1.86) | 1.72 (0.96, 3.09) | 1.13 (0.52, 2.48) | 0.65 (0.26, 1.62) |
| Antibiotics | 0.85 (0.67, 1.10) | 1.10 (0.70, 1.74) | 0.97 (0.54, 1.76) | 1.28 (0.68, 2.41) |
| csDMARD | 0.65 (0.42, 1.00) | 0.64 (0.28, 1.47) | 0.68 (0.22, 2.12) | 0.45 (0.09, 2.20) |
| bDMARD | 0.68 (0.37, 1.26) | 0.79 (0.25, 2.57) | 0.93 (0.17, 5.01) | 0.76 (0.09, 7.43) |
| bDMARD = biologics disease−modifying antirheumatic drugs; CI = confidence interval; csDMARD = conventional synthetic disease−modifying antirheumatic drugs; ICU = intensive care unit; OR = odds ratio; OS = overall survival.  Results are based on multiple logistic regression adjusting for age, race/ethnicity, sex, body mass index (BMI), comorbidities, and pre−index medication such as steroids, anti-coagulants, antibiotics, csDMARD and bDMARD. | | | | |

**Table S7	Demographics and Prior Medication Use by Hospitalization Among Patients with COVID−19**

|  | Non−Hospitalized  (n = 3107) | Hospitalized  (n = 949) | Overall  (n = 4056) |
| --- | --- | --- | --- |
| **Age** |  |  |  |
| Mean (SD) | 59.9 (16.2) | 66.8 (14.6) | 61.5 (16.1) |
| Median [Q1, Q3] | 62.0 [49.0, 71.0] | 69.0 [58.0, 78.0] | 63.0 [51.0, 73.0] |
| **Sex** |  |  |  |
| Female | 2554 (82.2%) | 697 (73.4%) | 3,251 (80.2%) |
| Male | 553 (17.8%) | 252 (26.6%) | 805 (19.8%) |
| **Race** |  |  |  |
| American Indian or Alaska Native | 11 (0.4%) | 8 (0.8%) | 19 (0.5%) |
| Asian | 51 (1.6%) | 10 (1.1%) | 61 (1.5%) |
| Black or African American | 825 (26.6%) | 330 (34.8%) | 1,155 (28.5%) |
| Native Hawaiian or Other Pacific Islander | 4 (0.1%) | 1 (0.1%) | 5 (0.1%) |
| White | 1603 (51.6%) | 423 (44.6%) | 2,026 (50.0%) |
| Unknown | 613 (19.7%) | 177 (18.7%) | 790 (19.5%) |
| **Ethnicity** |  |  |  |
| Hispanic or Latino | 381 (12.3%) | 140 (14.8%) | 521 (12.8%) |
| Not Hispanic or Latino | 1542 (49.6%) | 530 (55.8%) | 2,072 (51.1%) |
| Unknown | 1184 (38.1%) | 279 (29.4%) | 1,463 (36.1%) |
| **Smoking** |  |  |  |
| No | 2464 (79.3%) | 619 (65.2%) | 3,083 (76.0%) |
| Yes | 643 (20.7%) | 330 (34.8%) | 973 (24.0%) |
| **BMI (kg/m2)** |  |  |  |
| Mean (SD) | 28.6 (5.89) | 28.6 (6.17) | 28.6 (5.96) |
| Median [Q1, Q3] | 28.0 [24.0, 33.0] | 29.0 [25.0, 33.0] | 28.0 [24.0, 33.0] |
| Missing | 1930 (62.1%) | 553 (58.3%) | 2483 (61.2%) |
| **CCI (weighted score based on comorbid conditions)** |  |  |  |
| Mean (SD) | 1.50 (2.37) | 2.78 (3.08) | 1.80 (2.61) |
| Median [Q1, Q3] | 1.00 [0.00, 2.00] | 2.00 [0.00, 5.00] | 1.00 [0.00, 3.00] |
| **Steroids** ^a^ |  |  |  |
| No | 2,178 (70.1%) | 653 (68.8%) | 2,831 (69.8%) |
| Yes | 929 (29.9%) | 296 (31.2%) | 1,225 (30.2%) |
| **Anti−Coagulants** ^a^ |  |  |  |
| No | 2,961 (95.3%) | 853 (89.9%) | 3,814 (94.0%) |
| Yes | 146 (4.7%) | 96 (10.1%) | 242 (6.0%) |
| Any DMARD ^a^ |  |  |  |
| No | 2,718 (87.5%) | 854 (90.0%) | 3572 (88.1%) |
| Yes | 389 (12.5%) | 95 (10.0%) | 484 (11.9%) |
| **csDMARD** ^a^ |  |  |  |
| No | 2,804 (90.2%) | 876 (92.3%) | 3,680 (90.7%) |
| Yes | 303 (9.8%) | 73 (7.7%) | 376 (9.3%) |
| bDMARD = biologics disease−modifying antirheumatic drugs; BMI = body mass index; CCI = Charlson Comorbidity Index; csDMARD = conventional synthetic disease−modifying antirheumatic drugs; SD = standard deviation; tsDMARD = targeted synthetic disease−modifying antirheumatic drugs; Q1 = quartile 1; Q3 = quartile 3.  ^a^ All treatments mentioned were identified within a year of COVID−19 index date and prior to 2020. | | | |

**Table S7	Demographics and Prior Medication Use by Hospitalization Among Patients with COVID−19 (cont.)**

|  | Non−Hospitalized (n = 3107) | Hospitalized (n = 949) | Overall (n = 4056) |
| --- | --- | --- | --- |
| **tsDMARD** ^a^ |  |  |  |
| No | 3,075 (99.0%) | 940 (99.1%) | 4,015 (99.0%) |
| Yes | 32 (1.0%) | 9 (0.9%) | 41 (1.0%) |
| **bDMARD** ^a^ |  |  |  |
| No | 2,934 (94.4%) | 915 (96.4%) | 3,849 (94.9%) |
| Yes | 173 (5.6%) | 34 (3.6%) | 207 (5.1%) |
| bDMARD = biologics disease−modifying antirheumatic drugs; BMI = body mass index; CCI = Charlson Comorbidity Index; csDMARD = conventional synthetic disease−modifying antirheumatic drugs; SD = standard deviation; tsDMARD = targeted synthetic disease−modifying antirheumatic drugs; Q1 = quartile 1; Q3 = quartile 3.  ^a^ All treatments mentioned were identified within a year of COVID−19 index date and prior to 2020. | | | |

**Table S8	Demographics and Prior Medication Use by Death Among Patients with COVID−19**

|  | Survived (n = 3832) | Died (n = 224) | Overall (n = 4056) |
| --- | --- | --- | --- |
| **Age** |  |  |  |
| Mean (SD) | 60.8 (16.0) | 74.7 (9.68) | 61.5 (16.1) |
| Median [Q1, Q3] | 63.0 [50.0, 72.0] | 75.0 [68.8, 82.0] | 63.0 [51.0, 73.0] |
| **Sex** |  |  |  |
| Female | 3,096 (80.8%) | 155 (69.2%) | 3,251 (80.2%) |
| Male | 736 (19.2%) | 69 (30.8%) | 805 (19.8%) |
| **Race** |  |  |  |
| American Indian or Alaska Native | 17 (0.4%) | 2 (0.9%) | 19 (0.5%) |
| Asian | 60 (1.6%) | 1 (0.4%) | 61 (1.5%) |
| Black or African American | 1,091 (28.5%) | 64 (28.6%) | 1,155 (28.5%) |
| Native Hawaiian or Other Pacific Islander | 5 (0.1%) | 0 (0%) | 5 (0.1%) |
| White | 1,904 (49.7%) | 122 (54.5%) | 2,026 (50.0%) |
| Unknown | 755 (19.7%) | 35 (15.6%) | 790 (19.5%) |
| **Ethnicity** |  |  |  |
| Hispanic or Latino | 508 (13.3%) | 13 (5.8%) | 521 (12.8%) |
| Not Hispanic or Latino | 1,956 (51.0%) | 116 (51.8%) | 2,072 (51.1%) |
| Unknown | 1,368 (35.7%) | 95 (42.4%) | 1,463 (36.1%) |
| **Smoking** |  |  |  |
| No | 2,948 (76.9%) | 135 (60.3%) | 3,083 (76.0%) |
| Yes | 884 (23.1%) | 89 (39.7%) | 973 (24.0%) |
| **BMI (kg/m2)** |  |  |  |
| Mean (SD) | 28.6 (5.99) | 28.6 (5.58) | 28.6 (5.96) |
| Median [Q1, Q3] | 28.0 [24.0, 33.0] | 29.0 [25.0, 33.0] | 28.0 [24.0, 33.0] |
| Missing | 2,362 (61.6%) | 121 (54.0%) | 2,483 (61.2%) |
| **CCI (weighted score based on comorbid conditions)** |  |  |  |
| Mean (SD) | 1.67 (2.50) | 4.03 (3.32) | 1.80 (2.61) |
| Median [Q1, Q3] | 1.00 [0.00, 2.00] | 3.00 [1.00, 7.00] | 1.00 [0.00, 3.00] |
| **Steroids** ^a^ |  |  |  |
| No | 2,692 (70.3%) | 139 (62.1%) | 2,831 (69.8%) |
| Yes | 1140 (29.7%) | 85 (37.9%) | 1,225 (30.2%) |
| **Anti−coagulants** ^a^ |  |  |  |
| No | 3,625 (94.6%) | 189 (84.4%) | 3,814 (94.0%) |
| Yes | 207 (5.4%) | 35 (15.6%) | 242 (6.0%) |
| **Any DMARD ^a^** |  |  |  |
| No | 3372 (88.0%) | 200 (89.3%) | 3572 (88.1%) |
| Yes | 60 (12.0%) | 24 (10.7%) | 484 (11.9%) |
| **csDMARD** ^a^ |  |  |  |
| No | 3,471 (90.6%) | 209 (93.3%) | 3,680 (90.7%) |
| Yes | 361 (9.4%) | 15 (6.7%) | 376 (9.3%) |
| bDMARD = biologics disease−modifying antirheumatic drugs; BMI = body mass index; CCI = Charlson Comorbidity Index; csDMARD = conventional synthetic disease−modifying antirheumatic drugs; SD = standard deviation; tsDMARD = targeted synthetic disease−modifying antirheumatic drugs; Q1 = quartile 1; Q3 = quartile 3.  ^a^ All treatments mentioned were identified within a year of COVID−19 index date and prior to 2020. | | | |

**Table S8	Demographics and Prior Medication Use by Death Among Patients with COVID−19 (cont.)**

|  | Survived (n = 3832) | Died (n = 224) | Overall (n = 4056) |
| --- | --- | --- | --- |
| **tsDMARD** ^a^ |  |  |  |
| No | 3,794 (99.0%) | 221 (98.7%) | 4,015 (99.0%) |
| Yes | 38 (1.0%) | 3 (1.3%) | 41 (1.0%) |
| **bDMARD** ^a^ |  |  |  |
| No | 3,636 (94.9%) | 213 (95.1%) | 3,849 (94.9%) |
| Yes | 196 (5.1%) | 11 (4.9%) | 207 (5.1%) |
| bDMARD = biologics disease−modifying antirheumatic drugs; BMI = body mass index; CCI = Charlson Comorbidity Index; csDMARD = conventional synthetic disease−modifying antirheumatic drugs; SD = standard deviation; tsDMARD = targeted synthetic disease−modifying antirheumatic drugs; Q1 = quartile 1; Q3 = quatile 3.  ^a^ All treatments mentioned were identified within a year of COVID−19 index date and prior to 2020. | | | |

**Table S9	Demographics and Prior Medication Use by ICU Among Patients Hospitalized with COVID−19**

|  | Not Admitted to ICU (n = 809) | Admitted to ICU (n = 185) | Overall (n = 994) |
| --- | --- | --- | --- |
| **Age** |  |  |  |
| Mean (SD) | 63.8 (16.3) | 66.7 (13.3) | 64.3 (15.8) |
| Median [Q1, Q3] | 65.0 [55.0, 76.0] | 67.0 [60.0, 76.0] | 66.0 [56.0, 76.0] |
| **Sex** |  |  |  |
| Female | 614 (75.9%) | 126 (68.1%) | 740 (74.4%) |
| Male | 195 (24.1%) | 59 (31.9%) | 254 (25.6%) |
| **Race** |  |  |  |
| American Indian or Alaska Native | 5 (0.6%) | 0 (0%) | 5 (0.5%) |
| Asian | 6 (0.7%) | 0 (0%) | 6 (0.6%) |
| Black or African American | 281 (34.7%) | 91 (49.2%) | 372 (37.4%) |
| White | 362 (44.7%) | 85 (45.9%) | 447 (45.0%) |
| Unknown | 155 (19.2%) | 9 (4.9%) | 164 (16.5%) |
| **Ethnicity** |  |  |  |
| Hispanic or Latino | 78 (9.6%) | 8 (4.3%) | 86 (8.7%) |
| Not Hispanic or Latino | 477 (59.0%) | 125 (67.6%) | 602 (60.6%) |
| Unknown | 254 (31.4%) | 52 (28.1%) | 306 (30.8%) |
| **Smoking** |  |  |  |
| No | 516 (63.8%) | 107 (57.8%) | 623 (62.7%) |
| Yes | 293 (36.2%) | 78 (42.2%) | 371 (37.3%) |
| **BMI (kg/m2)** |  |  |  |
| Mean (SD) | 28.8 (6.28) | 28.4 (7.03) | 28.7 (6.42) |
| Median [Q1, Q3] | 29.0 [24.0, 33.0] | 29.0 [23.0, 33.0] | 29.0 [24.0, 33.0] |
| Missing | 437 (54.0%) | 98 (53.0%) | 535 (53.8%) |
| **CCI (weighted score based on comorbid conditions)** |  |  |  |
| Mean (SD) | 2.58 (3.02) | 3.22 (3.19) | 2.70 (3.06) |
| Median [Q1, Q3] | 1.00 [0.00, 4.00] | 2.00 [0.00, 6.00] | 2.00 [0.00, 4.00] |
| **Steroids** ^a^ |  |  |  |
| No | 544 (67.2%) | 123 (66.5%) | 667 (67.1%) |
| Yes | 265 (32.8%) | 62 (33.5%) | 327 (32.9%) |
| **Anti−Coagulants** ^a^ |  |  |  |
| No | 752 (93.0%) | 158 (85.4%) | 910 (91.5%) |
| Yes | 57 (7.0%) | 27 (14.6%) | 84 (8.5%) |
| **Any DMARD ^a^** |  |  |  |
| No | 725 (89.6%) | 166 (89.7%) | 891 (89.6%) |
| Yes | 84 (10.4%) | 19 (10.3%) | 103 (10.4%) |
| **csDMARD** ^a^ |  |  |  |
| No | 742 (91.7%) | 173 (93.5%) | 915 (92.1%) |
| Yes | 67 (8.3%) | 12 (6.5%) | 79 (7.9%) |
| bDMARD = biologics disease−modifying antirheumatic drugs; BMI = body mass index; CCI = Charlson Comorbidity Index; csDMARD = conventional synthetic disease−modifying antirheumatic drugs; ICU = intensive care unit; SD = standard deviation; tsDMARD = targeted synthetic disease−modifying antirheumatic drugs; Q1 = quartile 1; Q3 = quartile 3.  ^a^ All treatments mentioned were identified within a year prior to COVID−19 index date. | | | |

**Table S9	Demographics and Prior Medication Use by ICU Among Patients Hospitalized with COVID−19 (cont.)**

|  | Non−ICU  (n = 809) | ICU  (n = 185) | Overall  (n = 994) |
| --- | --- | --- | --- |
| **tsDMARD** ^a^ |  |  |  |
| No | 801 (99.0%) | 183 (98.9%) | 984 (99.0%) |
| Yes | 8 (1.0%) | 2 (1.1%) | 10 (1.0%) |
| **bDMARD** ^a^ |  |  |  |
| No | 780 (96.4%) | 176 (95.1%) | 956 (96.2%) |
| Yes | 29 (3.6%) | 9 (4.9%) | 38 (3.8%) |
| bDMARD = biologics disease−modifying antirheumatic drugs; BMI = body mass index; CCI = Charlson Comorbidity Index; csDMARD = conventional synthetic disease−modifying antirheumatic drugs; ICU = intensive care unit; SD = standard deviation; tsDMARD = targeted synthetic disease−modifying antirheumatic drugs; Q1 = quartile 1; Q3 = quartile 3.  ^a^ All treatments mentioned were identified within a year prior to COVID−19 index date. | | | |

**Table S10	Demographics and Prior Medication Use by Mortality Among Patients Hospitalized with COVID−19**

|  | Survived (n = 851) | Died (n = 143) | Overall (n = 994) |
| --- | --- | --- | --- |
| **Age** |  |  |  |
| Mean (SD) | 62.9 (16.1) | 72.8 (10.5) | 64.3 (15.8) |
| Median [Q1, Q3] | 64.0 [54.0, 74.0] | 73.0 [67.0, 80.0] | 66.0 [56.0, 76.0] |
| **Sex** |  |  |  |
| Female | 642 (75.4%) | 98 (68.5%) | 740 (74.4%) |
| Male | 209 (24.6%) | 45 (31.5%) | 254 (25.6%) |
| **Race** |  |  |  |
| American Indian or Alaska Native | 4 (0.5%) | 1 (0.7%) | 5 (0.5%) |
| Asian | 6 (0.7%) | 0 (0%) | 6 (0.6%) |
| Black or African American | 318 (37.4%) | 54 (37.8%) | 372 (37.4%) |
| White | 373 (43.8%) | 74 (51.7%) | 447 (45.0%) |
| Unknown | 150 (17.6%) | 14 (9.8%) | 164 (16.5%) |
| **Ethnicity** |  |  |  |
| Hispanic or Latino | 79 (9.3%) | 7 (4.9%) | 86 (8.7%) |
| Not Hispanic or Latino | 518 (60.9%) | 84 (58.7%) | 602 (60.6%) |
| Unknown | 254 (29.8%) | 52 (36.4%) | 306 (30.8%) |
| **Smoking** |  |  |  |
| No | 549 (64.5%) | 74 (51.7%) | 623 (62.7%) |
| Yes | 302 (35.5%) | 69 (48.3%) | 371 (37.3%) |
| **BMI (kg/m2)** |  |  |  |
| Mean (SD) | 28.6 (6.47) | 29.2 (6.17) | 28.7 (6.42) |
| Median [Q1, Q3] | 29.0 [24.0, 33.0] | 29.0 [25.0, 33.0] | 29.0 [24.0, 33.0] |
| Missing | 456 (53.6%) | 79 (55.2%) | 535 (53.8%) |
| **CCI (weighted score based on comobid contiions )** |  |  |  |
| Mean (SD) | 2.51 (2.99) | 3.81 (3.25) | 2.70 (3.06) |
| Median [Q1, Q3] | 1.00 [0.00, 4.00] | 3.00 [1.00, 6.50] | 2.00 [0.00, 4.00] |
| **Steroids** ^a^ |  |  |  |
| No | 576 (67.7%) | 91 (63.6%) | 667 (67.1%) |
| Yes | 275 (32.3%) | 52 (36.4%) | 327 (32.9%) |
| **Anti-coagulants** ^a^ |  |  |  |
| No | 787 (92.5%) | 123 (86.0%) | 910 (91.5%) |
| Yes | 64 (7.5%) | 20 (14.0%) | 84 (8.5%) |
| **Any DMARD** ^a^ |  |  |  |
| No | 763 (89.7%) | 128 (89.5%) | 891 (89.6%) |
| Yes | 88 (10.3%) | 15 (10.5%) | 103 (10.4%) |
| **csDMARD** ^a^ |  |  |  |
| No | 781 (91.8%) | 134 (93.7%) | 915 (92.1%) |
| Yes | 70 (8.2%) | 9 (6.3%) | 79 (7.9%) |
| **tsDMARD** ^a^ |  |  |  |
| No | 843 (99.1%) | 141 (98.6%) | 984 (99.0%) |
| Yes | 8 (0.9%) | 2 (1.4%) | 10 (1.0%) |
| **bDMARD** ^a^ |  |  |  |
| No | 821 (96.5%) | 135 (94.4%) | 956 (96.2%) |
| Yes | 30 (3.5%) | 8 (5.6%) | 38 (3.8%) |
| bDMARD = biologics disease−modifying antirheumatic drugs; CCI = Charlson Comorbidity Index; csDMARD = conventional synthetic disease−modifying antirheumatic drugs; SD = standard deviation; tsDMARD = targeted synthetic disease−modifying antirheumatic drugs; Q1 = quartile 1; Q3 = quartile 3.  ^a^ All treatments mentioned were identified within a year prior to COVID−19 index date. | | | |

**Figure S1. Standardized Mean Difference (SMD) of the matching variables pre and post matching for all COVID patients (solid blackline noted cut point of 0.1 SMD.SMD <0.1 is generally accepted as being balanced)**

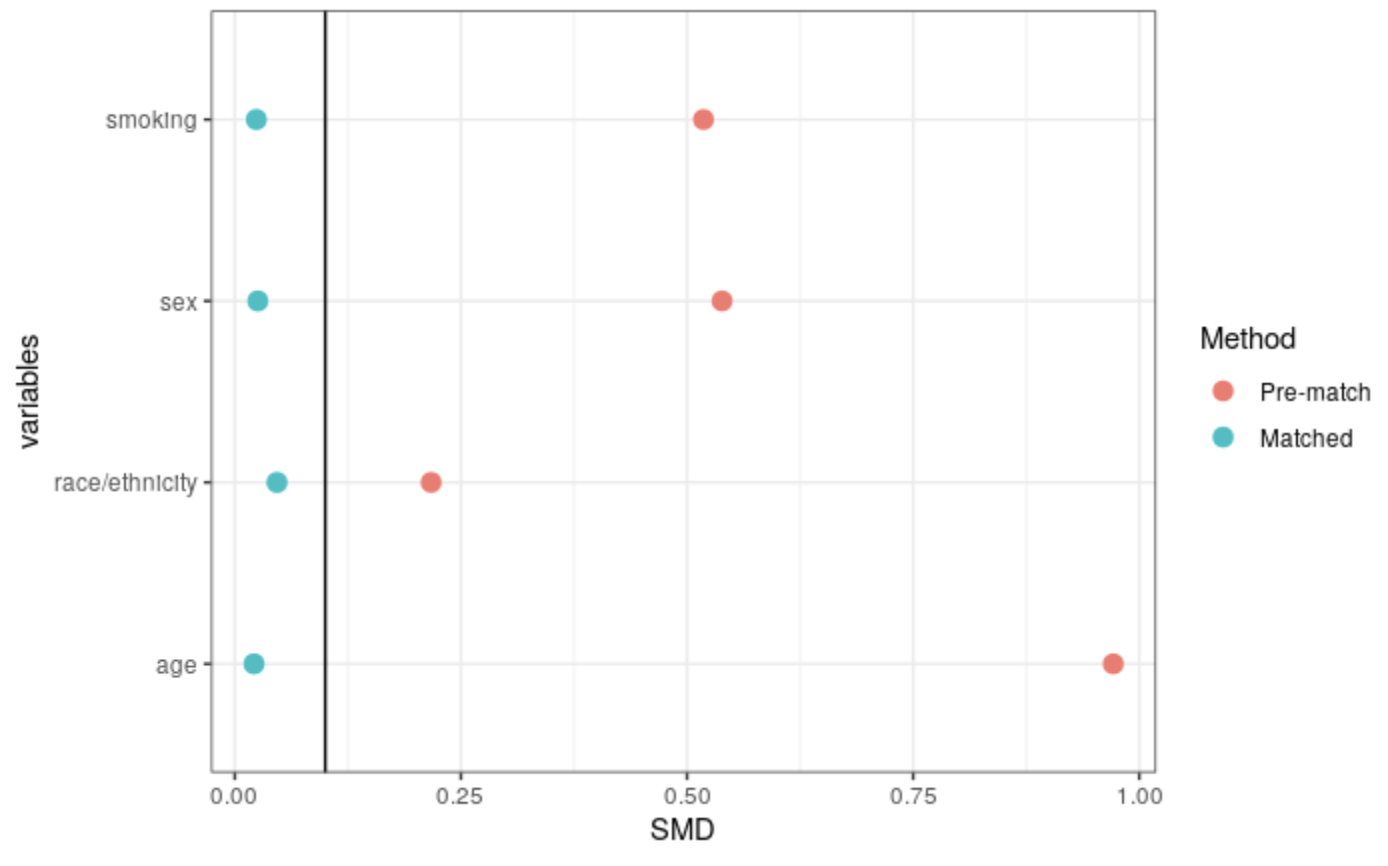

**Figure S2. Standardized Mean Difference (SMD) of the matching variables pre and post matching for hospitalized COVID patients (solid blackline noted cut point of 0.1 SMD.SMD <0.1 is generally accepted as being balanced).**

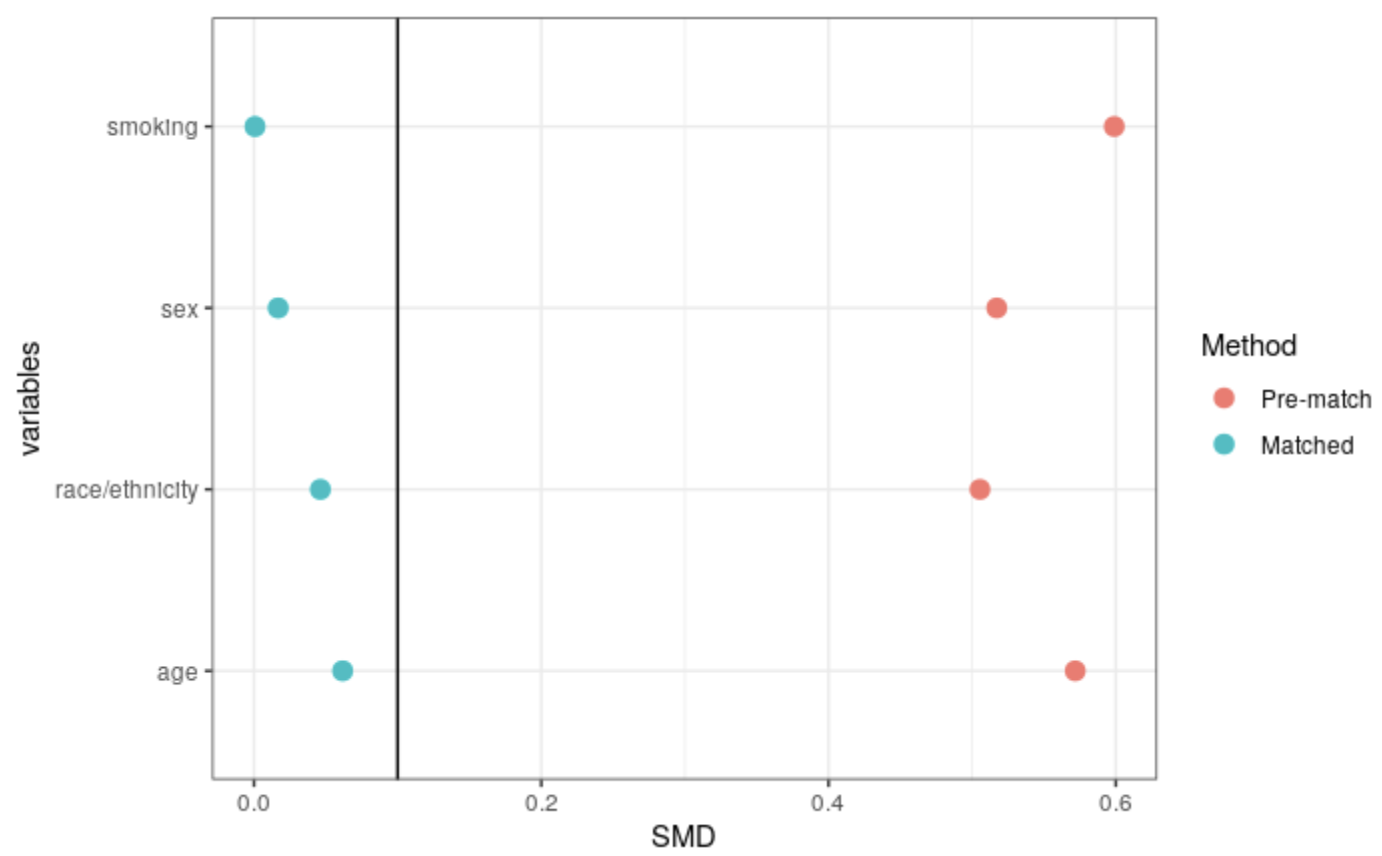
